# Barriers and facilitators to cross-sector workforce competency assessment in the care of older people residing in care homes

**DOI:** 10.1101/2021.01.21.21250213

**Authors:** Sue Tiplady, Juliana Thompson, Glenda Cook, Lesley Bainbridge

## Abstract

In recent years, transformation in care delivery towards cross-sector models has occurred with the aim of providing safe, efficient care for older people with complex needs, including those living in care homes. Effective workforce competency development and assessment across sectors is integral to these models. However, there is much evidence to suggest a distinct lack of cross-sector alignment with regard to competency development and assessment, which could lead to variations in practice and affect outcomes for older people. This qualitative study explored the barriers and facilitators to cross-sector workforce competency assessment in the care of older people residing in care homes. Twenty nine staff with responsibility for staff learning and development participated. Data were analysed using Braun and Clarke’s (2006) six phase thematic analysis approach. Findings indicate that a significant challenge to the development of a competent workforce is evidencing competency, due to difficulties in finding appropriate assessors and inconsistencies in the assessment process. Findings also indicate potential solutions including: a system-wide, standardised framework and strategy for assessment of competence; policy for the ‘sign-off’ of competencies that is recognised across organisational boundaries; and adopting practice-based approaches to competence development and assessment.

## Introduction

To maximise health outcome gains, the health and social care workforce, and the systems that influence workforce development, need to evolve to address changes in care contexts (World Health Organisation (WHO), 2015; 2016). Across the developed world, these changes include population ageing, and associated increasing rates of chronic disease, complex multi-morbidity, and frailty (AdvantAGE, 2018; Briggs, 2007; Marmot, 2014). In England, factors such as increasing patient expectations of care, health and social care workforce shortages, lack of desirability of working with older people, and the increasing intensity of care needs of care home residents add to the complexity of these changes (Buchan, Gershlick, Charlesworth, & Seccombe, 2019; Skills for Care, 2019).

Recognising that no one service provider can meet the increasing complexity of care needs, health policy has responded and adapted. NHS England (2020) recognises that being able to meet the complexity of care needs of individuals living in care homes can only be achieved by collaborative working between health, social care, voluntary, community, and care home partners. As such models of cross-sector working have emerged where professionals employed by one organisation will work into another with the aim of providing well-coordinated care for older people with complex needs (Cook et al.2016; NHS England, 2014; 2019; 2020; National Health Workforce Taskforce, 2010; Van der Heide et al., 2015). This cross-sector service provision is defined by Winter et al. (2016 p2) as ‘*independent, yet interconnected sectors working together to better meet the needs of consumers and improve the quality and effectiveness of service provision’*.

Care homes in England employ registered and unregistered care staff depending on the registered status of the home. Medical care for residents is provided by General Practitioners (GP), and further support is provided by a range of nursing and allied health professionals employed by the NHS or adult social services. Within England there are a reported 600,000 staff who work in care homes, with the largest proportion of staff employed in unregistered direct care-giving roles (Cavendish 2013; Skills for Care 2019).

Cavendish (2013) identified that all unregistered care workers should be equipped with fundamental and essential skills in the care of older people. This has contributed to the introduction and implementation of the Care Certificate, a skills development and assessment programme applicable to unregistered care staff working in all sectors (Skills for Care, 2015). Although the care certificate is not mandatory, there is an expectation that all staff new to health and social care will undertake the programme. The Care Quality Commission (CQC), the organisation responsible for regulating health and social care organisations, recognises the care certificate as good practice (CQC 2015). Recently, Dijkamen et al. (2016) identified a need for staff working in older people’s care to develop specific competencies related to the care of older people, regardless of the sector in which they work. Dijkamen et al.’s (2016) study subsequently contributed to the development of a European Core Competences Framework for Health and Social Care Professionals Working with Older People, further highlighting that workforce competence is integral to cross-sector working.

Although workforce competency frameworks and certificates exist that aim to provide competency standards in caring for older people (Bing-Jonsson, Bjork, Hofoss, Kirkevold, & Foss, 2014; Kiljunen, Partanen, Valimaki, & Kankkunen, 2018; Skills for Care, Skills for health & Health Education England 2015), these do not address the need for cross-sector working, nor offer guidance about standard competency assessment. Nevertheless, the relationship between cross-sector care outcomes and workforce competency is a topic of growing interest (Frenk et al., 2010; Hastings, Armitage, Mallinson, Jackson, & Suter, 2014; Langins, & Borgermans, 2015).

Despite aspirations of developing a workforce competent to deliver across-sectors, the reality of implementation remains challenging. A contributing factor to this challenge is that in the past, strengthening the workforce has focused on formal professional education programmes, when what is required are strategies to support and assess continuous competency development, maintenance and advancement in day-to-day practice over time (Barbazza, Langins, Kluge, & Tello, 2015; Levkovich, 2015). Even where competency development and assessment do occur, difficulties remain because strengthening competency across organisations requires system-wide change including strategic partnership working, alignment of operations, and competency development and assessment processes across all care sectors. However, there is much evidence to suggest a distinct lack of cross-organisational alignment and support, particularly between the health and social care sectors (Burger et al., 2018; Delaney, Robinson, & Chafetz, 2013; Kickbush, & Gleicher, 2012; Langins, and Borgermans, 2015; Levkovich, 2015).

This paper reports on one aspect of a wider study commissioned and funded by Newcastle Gateshead Clinical Commissioning Group, which aimed to understand the state of competency within a multi-sector workforce model caring for older people in care homes, located in North East England. This model is described in authors, 2017. This paper explores experiences and views regarding competency assessment of healthcare workers working within this multi-sector model. It also considers implications for the development of effective competency assessment practices for cross-organisational care systems.

## Background

In spite of the significance of competency to high quality care, and the introduction of competency frameworks, few studies consider assessment and validation of competency in older people’s care across care home and healthcare provider organisational boundaries.

Nicholson, et al.’s (2013) systematic review of integrated governance systems, and Barbazza et al.’s (2015) Europe-wide review of processes, tools and actors required for a competent workforce for cross-sector care, highlight that accredited competency development programmes with commonly agreed standards and assessment processes is essential. While these reviews give examples of innovations in developing accredited programmes, little is said about how the competencies of individuals could be effectively assessed. Kiljunen et al.’s (2019) study provides insight into the challenges of using self-assessment as a measure of competence of Finnish care home nurses, identifying differences between self-assessed and manager-assessed competency, but the study did not consider cross-sector assessment. Thomson et al.’s (2018) evaluation of The Care Certificate does not investigate cross-sector assessment, but does highlight that there are a number of inconsistencies across organisations regarding how the care certificate is implemented including variations in the delivery of training and how staff’s competence is assessed. Thomson et al. (2018) concludes that inconsistencies between organisations has undermined the credibility and portability of the Care Certificate. Varkey, Reller, & Rasar’s (2013) review of quality improvement tools suggests audits and feedback can be used to identify gaps and variations in competencies. Whilst this study demonstrates the use of improvement tools in competence assessment, it does not make clear how these relate to competency assessment of individuals working in cross-organisational settings. Lockyer’s (2003) study proposes that multi-source or 360-degree evaluations are effective as a means of assessing competency in interpersonal and communication skills, professionalism, and systems-based practice, but found they are less suitable for clinical and practice-based competency assessment.

Other studies, while not considering care homes, investigate assessment across different sites, countries, management structures and cohort positions, which provide some useful insight into competency assessment across organisational boundaries. Palermo et al.’s (2015) Australian study of dietician graduates’ experiences of competency assessment suggests assessment in work-based settings are subjective and inconsistent primarily because of variations in standards across sites. These authors conclude that this, together with lack of clarity about what a competent practitioner looks like, and a dearth of appropriately trained supervisors and assessors, leads to difficulties in creating appropriate learning opportunities and fair assessment of competency. Studies undertaken in Saudi Arabia, where 68% of nurses are recruited overseas, suggest that because different countries have differing competency frameworks and competency assessment procedures, employers with large international health and social care workforces are presented with difficulties in achieving consistent standards, and safe quality care (AlYami, & Watson, 2014; Colet et al, 2015; Salah et al., 2017). Salah et al. (2017) conclude that where a workforce is drawn from many countries, then standardisation of competency validation is essential to ensure all health and social care professionals meet expected and required competencies.

Cusack and Smith (2010) propose that competency assessment can be problematic where there are perceived power inequalities resulting from complex workforce environments, professional hierarchies, or healthcare cultures. This study builds on the seminal work of Thompson (1989) who identified that social expectations based upon deep-seated cultural traditions, and social/political tensions between different groups are at play within work environments, which can influence work practices. Where such tensions are apparent, ‘hostile’ work environments can ensue. According to Cusack and Smith (2010), hostile work environments in healthcare, resulting from professional hierarchies, and differences between workers’ cultural and professional situations and backgrounds, can hinder effective competency assessment. For example, assessors may feel anxious or uncomfortable about assessing workers with differing backgrounds to their own, or not want to assess others working in different organisations. On the other hand, assessors may be judgemental and critical of assessees from other backgrounds, devaluing their skills and knowledge, which can have a detrimental impact on assessees’ self-esteem and confidence. Thompson et al.’s (2016) study of the occupational role and status of care home nurses suggests that care home nurses’ perception that NHS staff do not value their contribution to care for older people can lead to mistrust and conflict between these staff groups.

While the literature to-date provides some insight into the difficulties encountered in competency assessment across settings, further research is required to explore competency assessment in the context of cross-sector working regarding care homes for older people with complex needs.

## Method

COREQ guidelines (Tong, Sainsbury & Craig, 2007) were used in reporting this study.

This study was undertaken by researchers experienced in gerontological research and workforce development research. As the study explored barriers and facilitators to cross system workforce competency development and assessment, a qualitative methodology within a constructivist paradigm was adopted. This approach is appropriate for exploration of shared meanings and understandings within organisational and policy contexts (Crotty, 1998).

Research ethics approval to undertake the study was secured from the Faculty of Health and Life Sciences, Northumbria University.

### Sample

Professionals working within a multi-sector workforce model caring for older people in care homes, located in North East England, and who had responsibility for staff learning and assessment were identified from university continuing professional development and from local Clinical Commissioning Group (CCG) records (n=278). These individuals were sent an email inviting them to participate in the study. In total 29 individuals responded and consented to participate (response rate of 10.4%). Following consent to participate in the study, all respondees took part in an interview. Participants worked in 6 private care home organisations, one local authority funded care home, and National Health Service (NHS) organisations.

**Table 1:**
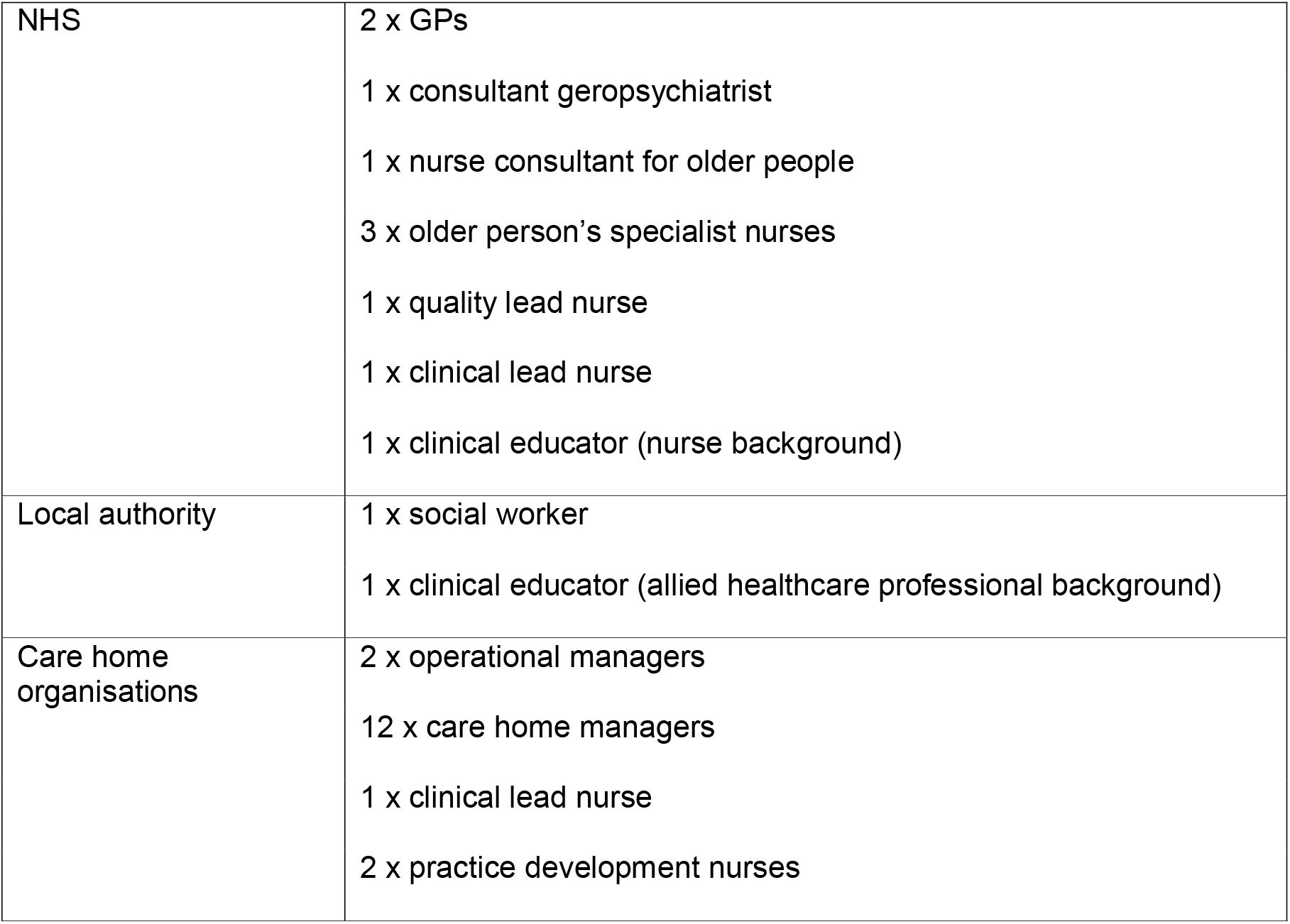
Study participants.

|  |  |
| --- | --- |
| NHS | 2 x GPs<br>1 x consultant geropsychiatrist<br>1 x nurse consultant for older people<br>3 x older person's specialist nurses<br>1 x quality lead nurse<br>1 x clinical lead nurse<br>1 x clinical educator (nurse background) |
| Local authority | 1 x social worker<br>1 x clinical educator (allied healthcare professional background) |
| Care home organisations | 2 x operational managers<br>12 x care home managers<br>1 x clinical lead nurse<br>2 x practice development nurses |

In order to maximise confidentiality, when reporting data in the form of participants’ verbatim quotes, their employing organisations only are given.

### Data collection

The planned data collection approach was focus group interviews. However, not all staff agreeing to participate could attend group interviews. The research team therefore adopted a pragmatic approach to maximise participation, by utilising a mixture of focus group, dyad and individual interviews. This approach maximised opportunities for practitioners to participate. Data was collected through 2 focus group interviews (n=9 and n=10), 2 dyads and 6 individual interviews. For participants’ convenience, data collection took place at their work place, and lasted for no more than one hour. During the interviews, participants were asked to discuss, and give examples of, barriers and facilitators to competency development and assessment; what preparation and systems are required to enable competency development and assessment, and what are the implications of competency development and assessment for staff, patients/service users, and organisations. Participants were asked to consider these topics within an integrated care model or working across organisations and sectors.

### Data analysis

Audio recordings were made of the focus groups, dyads and individual interviews. The audio-recorded data was transcribed verbatim, and open coded by individual members of the research team. This allowed elucidation and description of participants’ experiences of competency assessment, while creating meaningful themes. Thematic analysis was chosen as it is ‘a method for organising, analysing and reporting patterns (themes) within data. It minimally organises and describes your data set in (rich) detail’ (Braun and Clarke, 2006, p.79). The approach taken was inductive, in other words the analysis was data driven, rather than theory driven. The 6 phase guide to conducting thematic analysis, as outlined by Braun and Clarke (2006) was used. This process has the following phases: familiarisation with the data; generating initial codes; organisation of initial codes into patterns to generate themes; reviewing themes; defining and naming themes; interpretation. During this process, all transcripts were independently coded by another team member, and outcomes were compared with the original coding to validate themes.

## Findings

Participants recognised that learning, development and the assessment of competence were important and beneficial for older people, staff, the wider health and social care system and care home providers:

> The residents benefit totally. If your nurses are competent and your carers are competent, they are the ones that benefit from it. You know the care is good. So, at the end of the day, it’s the residents that benefit from it. Plus the staff, because they feel proud in themselves that they’ve learnt to upgrade their skills (Care Home participant).

However, most participants proposed that competency assessment needs to be consistent and standardised across health and social care sectors and organisations to ensure older people, who access care are provided with safe, effective care:

> I think everybody would accept - or most people would accept and understand - that with standardisation comes improved safety. And therefore improved efficiency and care delivery. And that has to bring system benefits. (NHS participant).

Respondents indicated that a significant challenge concerning the development of a competent workforce is demonstrating competency, due to difficulties in carrying out competency assessment. Two themes regarding this potential challenge emerged from the data analysis: barriers and facilitators to finding appropriate assessors, and barriers and facilitators to the assessment process.

### Barriers and facilitators to finding appropriate assessors

Some participants suggested that while education establishments provide development opportunities, they do not routinely offer competence assessment as part of educational programmes. For example, a range of learning opportunities could be accessed by care home staff, however much of this provision focused on knowledge development without assessing how that knowledge was applied in practice. Determination of competence to practice was left as a responsibility for the employing organisation:

> The nurses can go on a ten-day clinical course over three months. They’ll do the theory. They’ll do a practice session on that day. For example, doing a catheterisation, they’ll use a model where you insert the catheter. They’ll monitor you doing that, but then they give you a competency framework to take away, to be signed off by staff that are in your home (Care home participant).

This suggests that education providers acknowledge that academic or knowledge-based study on its own is not an indicator of clinical competence, and that work-based supervised practice is integral to competency assessment.

Some participants identified the requirement for assessors of clinical competencies to be competent themselves as clinicians with expertise in the care of older people. Participants said that often, the most experienced clinicians were promoted into management positions resulting in a move away from clinical practice. Participants felt this compromised these clinicians’ ability to assess competency:

> Years ago, I did my teaching certificate and my mentorship certificate. But, of late, because I don’t work physically on the floor, there are times when I’ll say, ‘No, I can’t assess your competency because I haven’t done it myself, for a while’ (Care Home participant)

Many care homes employ non-nurse managers so do not have nurses with specific knowledge of caring for older people, or indeed, do not have a stable registered nurse staff base. This means that access to competence assessment in house can be limited:

> A lot of the managers are non-nurse managers. So, they’re not able to sign them off. And a lot of the homes don’t have a cohort of nurses, so they’re relying on agency staff. So, there is an issue as to who’s deemed them as competent. What’s happened in the past is I’ve had phone calls, because I’m one of the very few nurse managers, being asked can I go along and assess their competency? Or, can my clinical leads that I have here go across and assess their competencies for them? (Care home participant)

Within this company, this manager or her staff often assessed the competency of staff in other homes, which led to depletion of service in their own care home.

An alternative approach to assessment of competencies in the care home sector could be other professionals working in sectors such as the local authority or NHS fulfilling this function. However, in most cases, participants said that cross-organisational competency assessment was problematic because assessors were concerned about implications concerning accountability arising from assessing staff from another organisation. In the cases below, participants were discussing nursing staff assessing other nurses across-sectors:

> Because they’re employed from a different organisation to us. So, I have asked them to assist, but because they’re employed by another organisation company, they don’t assist, and can’t assist (Care home participant).
>
> So, this is a bit dodgy…You know, like there are issues of accountability (NHS participant).

A further challenge arose with regard to the possibility of cross-profession assessment. Some participants indicated that medical staff were well positioned to assess nursing and allied healthcare professionals as they work closely with them. They also provide informal training while discussing the care of patients:

> But informally I think we do provide, like, support staff in terms of training and just, like, informally in conversation when reviewing patients’ (NHS participant)

However, some medical participants stated that they did not feel able to assess individuals from other professions because they do not have enough knowledge about the competency assessment processes of other professions:

> I’ve got my comfort zone in terms of I know what I’m doing with medical students and junior doctors, but the thought of trying to assess a competency in a nurse, an AHP, a care home professional, in a formalised sense, is quite anxiety-provoking to me (NHS participant).

Other medical staff participants expressed willingness to assess competence but acknowledged the need for preparation and guidance to assess competence across professional groups:

> I’ve been trained to train and to assess their [doctors’] competency. But I probably would need some guidance from a nursing point of view. Like, what would be expected or… how to do it formally (NHS participant).

Being assessed by individuals working for external agencies was also of concern due to perceived tensions in relationships between health and social care organisations, but also because the assessee may have little knowledge about the assessors’ own competency levels:

> Historically, the relationship between health and social care hasn’t been the best. There’s always been that hierarchical attitude in my view, of NHS staff coming into care homes. And that Cinderella service - it’s still not brushed off. And so, I think there would be a reluctance within the care homes to be assessed by those people. But, actually, we don’t know what their competencies and skills are (Care home participant).

One participant explained how a local care home has successfully addressed the problem of cross-organisational competency assessment. In this case, the local authority employed a workforce development officer with a healthcare education background. This person works with both the NHS professional providing support to the care home staff, and the care home staff themselves, and in tripartite meetings, supports the NHS professional through the competency assessment sign off process. As this person has independent knowledge of the NHS professional’s clinical competency, and is involved in the sign-off process thereby sharing accountability with the NHS professional, the system is acceptable to all parties:

> The other thing you’ve got to think about is who signs off the competency when you’re talking about the sign off from a different organisation. We overcome that by involving the workforce development officer from the local authority (Local Authority participant)

### Barriers and facilitators to the assessment process

Some participants identified issues with cross organisation assessment of competence:

> There were a lot of barriers to assessing nurses who work in different organisations - there’s a big organisation block, because they’re frightened of allowing nurses working in one organisation to assess in others… For example, nurses working in an NHS organisation assessing the competence of care home staff. I think that scares them. (NHS participant). This participant further identified that there was a need for robust governance measures to be in place to mitigate against cross-sector concerns.

Lack of consistency in competency assessment also occurred as a result of assessors’ different views about what is required to evidence competency. For example, in the following case, the participant felt that clinical activities should be observed 6 times prior to competency sign-off to ensure effective competency assessment:

> I don’t see them on a regular basis, doing it. I’m signing somebody off, but I’m not watching them in practice all the time. I’m just seeing that one-off session. If it’s venepuncture, I would have to observe them 6 times. But, I’m only seeing a snapshot (NHS participant).

While this participant said that clinical activities had to be observed 6 times prior to competency sign-off, this was not consistent across all organisations. Some participants reported that 3 observations were sufficient for sign-off, and others suggested up to 10 observations. Defining the number of repeated observations required is an attempt to improve the validity of assessment to ensure the person is proficient in different contexts and situations, however there is variation in the number of observation episodes required to make an accountable decision regarding proficiency.

Many participants proposed that some aspects of practice are difficult to assess, such as competency assessment of values:

> Certainly all the values stuff is very difficult to assess. Particularly if you’re trying to put it into a… quantitative way, but into a formalised sense of what constitutes good enough values, appropriate values (NHS participant)

Another aspect of practice that some participants felt is difficult to assess is comprehensive, holistic care of older people living with multimorbidity and complex care needs:

> Saying holistic - but it’s very easy to say can someone put a catheter in? Can someone do a respiratory exam or a cardiac exam? But, actually, when you’ve got these complex people that have got COPD and heart failure and diabetes, it’s how do you assess whether someone can manage that complexity and multi-morbidity (NHS participant).

A further barrier to competency assessment reported by participants was the time required to assess competence and the need for this to be a core aspect of practice rather than an added extra in one-off pre-determined situations:

> As I said before, it needs to be part of their daily practice, and not additional to their daily practice. Because nobody has got time to do anything extra (NHS participant).

Participants suggested solutions to these competency assessment challenges. One solution that was considered particularly successful was the use of CCG funded clinical educators. NHS clinical educators were seconded into a care home support role where they provided competency development activities and carried out competency assessment. These clinical educators devised some observational activities and questionnaires as a method of assessing competency in practice, which they used as standard across all care homes. This approach enabled assessment of all aspects of practice including values, and complex care. Assessment occurred during practice hours, which was time efficient, but also ensured assessment was relevant and meaningful:

> To try and standardise the observations, we had standardised questions. So, it would be a case of, what you would expect to see. And if you didn’t see that, what questions you might ask to reinforce it. So, you might see somebody walking along and prompting somebody to use the walking frame properly. But you wouldn’t particularly see them checking the safety features in it. So, you would ask, ‘What are the safety features?’ So, it’s just trying to standardise what you want to see to be signed by somebody as competent (Local Authority participant).

In this example, validity and reliability of the assessment process was enhanced in a number of ways. Rather than relying solely on observations of practice, assessors built a series of questions into the assessment process to check underpinning knowledge behind practice activities. This meant that assessment could extend beyond observed care episodes. Also, assessors were skilled in the care of older people, as well as having expertise in workforce development. In addition, a standardised assessment framework was used so that assessment against agreed criteria could be facilitated.

## Discussion

As the population continues to age, numbers of older people with complex needs including multimorbidities, disability, severe frailty and terminal illness will rise, increasing demand on health and social care services. This, together with workforce shortages across health and social care has been identified as adversely affecting older people’s access to services and increasing their risk of hospitalisation (Age UK 2019; Care Quality Commission, 2019). This reinforces the requirement for a workforce that is efficient, effective and competent in the delivery of enhanced care for older people (Cavendish, 2013; Langins, & Borgermans 2015; Burger et al. 2018).

When discussing the challenges that are currently faced regarding workforce and competency development, participants were in favour of adopting a standardised approach competency framework across health and social care systems, and highlighted that this could improve safety and efficiency. This reflects findings from Cavendish Report (2013), and Langins and Borgermans’ report for WHO (2015) whereby care organisations identified they would welcome an agreed set of national competences for staff, stating these would help improve safety, raise care standards and potentially be cost effective.

However, findings indicate that this on its own is not an adequate workforce development strategy. Despite the value of standardised frameworks being acknowledged, participants discussed difficulties in meeting required competency standards due to challenges in achieving standardised, effective competency assessment. A major concern for participants was how competence should be assessed. Findings highlighted that inconsistencies in assessment lead to difficulties in making conclusions about staff competence, and can result in mistrust between staff working in different sectors and organisations these findings were mirrored in the evaluation study of the care certificate (Thomson et al 2018). Langins and Borgermans (2015) suggest that, with no gold standard being available for measuring clinical competence in the field of older people’s care, there is a need for a system level strategy for agreeing required outcomes, standards of performance and competence assessment. Barbazza et al. (2015) highlight that critical to the successful development of a competent workforce are robust governance arrangements that support competency development and assessment across-sectors. Findings from this study suggest similar solutions when discussing the challenges and facilitators to competency assessment processes. These include the need for a system-wide, standardised framework for assessment of competence to go alongside the competency framework, and a need to adopt practice-based approaches to competence assessment to maximise efficiency and ensure assessment is relevant and meaningful to practice.

A further concern for participants was finding appropriate assessors - a difficulty stemming from obstacles to cross-organisational working and support. This resonates with studies by Palermo et al. (2015), Burger et al. (2018) and Langins and Borgermans (2015) that refer to the considerable debate about who should assess competence and in what circumstances, and the issues about governance and liability with regard to who can ‘sign off’ competence to practice. Findings from this study identified staff’s reluctance to assess across organisational and professional boundaries due to accountability concerns, lack of understanding about expected competency levels of other professions, and tensions between sectors. This is similar to Cusack and Smith’s (2010) research that identified differences in professional situations can lead to ‘hostile’ work environments which can hinder effective competency assessment. Participants in this study were also concerned that assessors should be clinically credible themselves. This reflects Wyman et al’s (2019) study of the development of a recognition programme for educators of gerontological nursing, which concludes that educators in gerontological healthcare need to be competent in this area of practice.

The study findings indicate methods of addressing these challenges, namely by the employment of clinical educators who can work across-sectors using a standard, practice-based assessment of competency approach, including competency ‘sign-off’, that is recognised across-sector and organisational boundaries. Consequent to the wider study, of which the findings reported in this paper are a part, the local CCG led a successful application for funding to employ two full-time equivalent Strategic Workforce Development Leads for Older People. These postholders will lead, support and facilitate workforce competency development across the region to enable those caring for older people to meet the required competencies identified within the standardised workforce competency framework adopted by the CCG, by building cross system infrastructure and capacity for practice based learning and assessment. They will use a cascade model of competency development; firstly developing a standardised development and assessment framework aligned with the competency framework standards, then they will identify individuals with expertise in the care of older people from across the region to train as practice educators and assessors. This staff group will then work in practice, across-sectors to support and assess competency development. Adoption of this model will facilitate the introduction of a ‘passport of competence’ which staff with competence aligned to the competency framework will hold. This initiative, originally recommended by Cavendish for the unregulated workforce (2013) and reinforced by Laycock, Borrows and Dobson (2017) in a report for reform as a means of standardising and evidencing competence of health care assistants so competence is accepted universally across-sectors.

### Limitations of this study

This study’s findings are based upon the responses of a small number of participants located in one location in England. It is recognised that findings may reflect local challenges and solutions. However, the challenges identified in the study location resonate with findings from other national and international literature, suggesting a degree of transferability. The recommendations suggested in this study, namely the introduction of Strategic Workforce Development Leads and ‘passports of competence’ require evaluation subsequent to their implementation to assess their effectiveness and impact on practice outcomes.

## Conclusion

Workforce competency development is integral to health and social care systems that aim to be closely aligned or integrated. There are significant challenges concerning the demonstration of staff proficiency in specific competencies, due to difficulties in finding appropriate assessors and inconsistencies in the assessment process. Adoption of a system-wide, standardised framework for assessment of competence alongside a competency framework is one solution to address this issue. This approach can enhance inter-professional working and support, enhance workforce competence, and ultimately improve care for older people. In the study location, this is now being achieved via the employment of Strategic Workforce Development Leads for Older People to build cross system infrastructure and capacity for practice based learning and assessment; and the adoption of a ‘Passport of competence’ which staff with competence aligned to a standardised framework will hold.

## Data Availability

Data is available by contacting the authors.

## Acknowledgements

The authors would like to thank Newcastle Gateshead Clinical Commissioning Group for commissioning and funding this research.

